# Quantifying infection-relevant contact patterns among young children in childcare settings in the United States

**DOI:** 10.64898/2026.08.18.26360623

**Authors:** Sara L Loo, Anjalika Nande, Shaun Truelove, Alison L Hill

**Author notes:** Corresponding author, 25 Willcocks Street, Toronto, Ontario, Canada M5S 3B2. These authors contributed equally to this work (Sara L Loo, Anjalika Nande).

## Abstract

Age is a primary determinant of symptom severity and transmission patterns for many infectious diseases, motivating the use of age-stratified models parameterized by contact matrices. In the United States, the absence of direct contact surveys has required estimating synthetic contact matrices from demographic data on household size, school attendance, and workforce participation. However, this likely underestimates contacts among children under age 5, who often attend group childcare missing from censuses. The goal of this study was to use nationally-representative data on childcare arrangements (the Early Childhood Program Participation Survey) to reconstruct daily contacts occurring in childcare settings, and augment existing all-age contact matrices. For infants under 1 year of age, we estimated 0.2 daily contacts with other infants, increasing to 0.7 daily contacts with same-age peers for 1- or 2-year-olds, 1.3 for 3-year-olds, and 3.5 for 4-year-olds. Including childcare settings increases estimated contacts among young children by up to six fold. Using simulations of measles outbreaks in inadequately vaccinated populations, we show that prior contact matrices significantly underestimated the outbreak frequency, size, and impact on preschool age groups. Our findings highlight the need for targeted data collection on childcare contacts to improve model-based evaluation of interventions particularly for young children.

## Introduction

For many infectious diseases, age is a primary determinant of symptom severity and of the patterns of physical contact that can lead to transmission. Thus, classical compartmental mathematical models of infectious disease spread often stratify individuals by age, and describe the rate of disease transmission between different age groups with a contact matrix. Contact matrices and person-to-person interactions are notoriously difficult to measure, but typically describe the daily number (or frequency) of contacts meeting certain spatial and temporal criteria and have been estimated in the past using physical proximity sensors (1–3) or mobile phone data (4, 5), with detailed individual-level surveys or contact “diaries” (6–11), approximated with data from existing demographic censuses (12–14), or inferred from patterns of disease spread (15, 16). These matrices are often composed of several setting-specific matrices, for example contacts in households, workplaces, schools, or the general community.

The burden of many infectious diseases is concentrated in young children. American children under the age of 5 on average have 4 - 7 upper respiratory infections a year (17). Pediatric infections have significant direct and indirect economic costs for households, governments, and other payers (18). For example, in the United States, hospitalizations due to respiratory syncytial virus cost approximately $124 per all births per year (19), and before vaccines were introduced, hospitalization with rotavirus infection was estimated to lead to 3.4 lost days of work for parents and a median direct cost of $4565 per hospitalization (20). The more severe outcomes of infections in young children are often attributed to relative lack of immunity (due to waning maternally-acquired antibodies, lack of prior exposure, and vaccination schedules) or other physiological characteristics. The role of increased transmission due to high rates of close physical contact is a common anecdotal explanation but has rarely been formally evaluated. Contacts among children may be especially poorly captured in existing contact matrices. Even with detailed contact diary surveys, responses for younger children are recorded by parent proxies who are likely not present with children in school, and problems of “recall” are likely to be especially salient.

In the United States, while there have been some recent state-level surveys and studies investigating contacts, and particularly changes in contacts during the COVID-19 pandemic (e.g. 7, 21), there is a lack of comprehensive (geographically and age-specific) survey data capturing age-by-age specific contacts relevant to infections spread by close contact, of the type collected in the POLYMOD study covering 8 European countries (6) or in the GlobalMix study covering 4 LMIC countries (22). Instead, two highly cited papers have attempted to impute a plausible contact matrix by using high-level census data and survey derived contact matrices from other countries (12, 13). These contact matrices are the primary sources used in US-based mathematical models (for e.g., 23–29), but they suffer from a serious missing data problem for child-to-child contacts among the youngest children. In the vast majority of US localities, public schooling begins between ages 5 and 6, at “kindergarten”. Before that, however, most young children and even many infants are in childcare while their guardians work (30), split roughly between home-based care (which is often informal and unlicensed) and more formal “center-based” care. In these childcare settings, children are often in very close contact with other non-sibling, or non-household, children. However, for the most part, information on childcare arrangements is not recorded in the census or other datasets (which are used to inform synthetic contact matrices (13)). Respondents can indicate “preschool or nursery school” as an option for individuals age 3 and older. However, these terms are not clearly defined, and do not reflect the most common American terms for childcare; e.g., an unlicensed neighbor caring for three unrelated children, would typically be considered home daycare, not a “preschool”. Similarly, UNESCO data used in Prem et al. (12) does not include informal care. This limited information misses children under three years old, and even for ages 3-5, fails to capture the diversity of childcare settings and contact patterns, resulting in lower than expected average daily contacts; e.g., the average number of daily contacts between 3 year olds is approximately 1.175, while between 2 year olds it is even lower, at 0.178.

In this study we use data from an in-depth national survey of childcare arrangements for children in the United States to reconstruct realistic age-stratified contacts for young children, and augment existing matrices to be used in infectious disease models (Figure 1). The Early Childhood Program Participation (ECPP) Survey is administered by the National Center for Education Statistics (NCES) every few years to a random subset of guardians of young children (identified via census responses) (31). It collects detailed data on the type(s) of childcare used including the setting, provider type, and hours of attendance, in addition to characteristics of the child and their household. We use the results from this survey along with other sources such as federal guidelines governing licensed childcare facilities to generate synthetic age-structured childcare settings.

**Figure 1:**
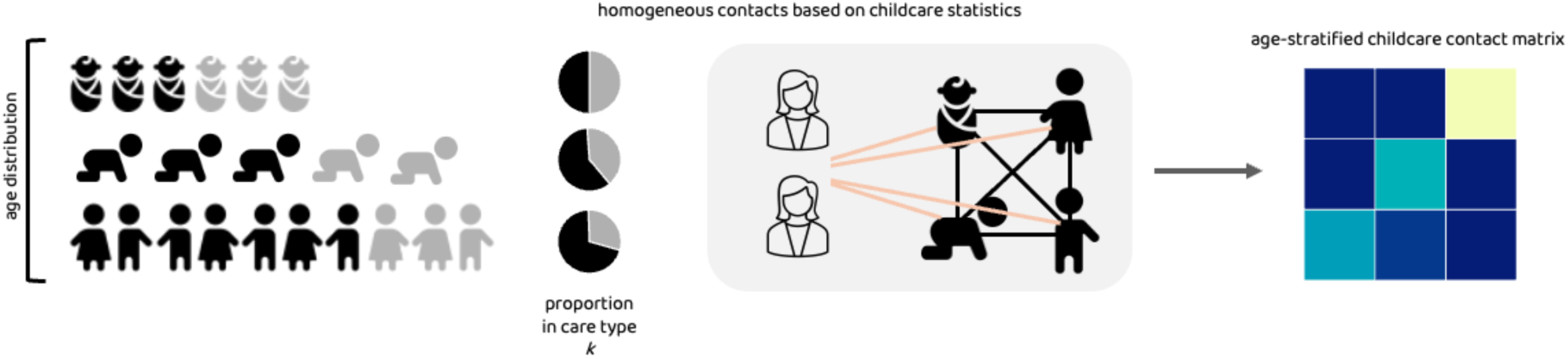
Overview of the construction of contact matrices in childcare settings. Our approach combines information on the age distribution of children under 5, the proportion in each age group in various care types and settings, the frequency of attendance in care, and characteristics of the care setting including class sizes, age groups, and caregiver ratios. We create synthetic childcare settings with statistics matching reported values, and assuming homogeneous mixing within a childcare setting on a given day, create matrices estimating daily contacts per age-by-age group pair.

From this, we estimate age-by-age average daily contacts for children from infancy through 5 years at a monthly age resolution to produce updated contact matrices to input into transmission models. Our findings suggest that contacts among young children have been dramatically underestimated by prior synthetic contact matrices for the US. Consequently, prior modeling studies using these matrices have likely underestimated the predicted contribution of young children to population-level disease transmission and the importance of disease control in these groups. To highlight the relevance of these results, we demonstrate the effect of our augmented contact matrix on the predicted impact of measles introductions into four scenarios of changing vaccination coverage in children. Our results stress the need to develop more targeted surveys to quantify contacts in young children, especially given the diverse childcare arrangements prevalent in the United States.

## Methods

### Modeling contacts in childcare between young children

Our goal is to calculate the contact matrix **C** whose values *C_i,j_* are defined as the average number of contacts per day that a child of age *i* (*index*) has with children of age *j* (*contact*) due to interactions in childcare environments. Following the methodology of the ECPP survey (31), we divide childcare setting types *k* into i) in-home care by a relative caregiver, ii) in-home care by a non-relative caregiver, and iii) center-based care. The overall contact matrix can be decomposed into contributions from each care type,

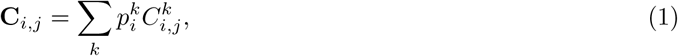

where 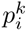 is the proportion of children in age group *i* who receive care in care type *k* on any given day, 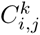 is the average number of contacts per day that a child of age *i* has with children of age *j* due to interactions in childcare type *k*. Note that children can attend more than one type of care so we can have 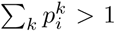. Our notation is summarized in Table S1.

Considering the simplest scenario where we assume that children mix homogeneously with all other children receiving care in the same care type:

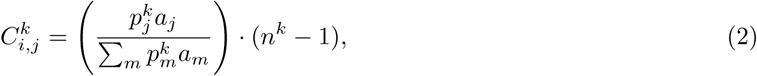

where *a_j_* is the fraction of the entire population in age group *j* and *n_k_* is the average number of children being cared for together in care type *k*.

We note that 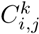 is independent of *i*, and is given by the age distribution of children in that childcare type in general. This is a reasonable assumption for home care arrangements where there are fewer children and a smaller likelihood of being divided into “classes”. For those in center-based care, we assume that children are instead divided into “classes” based on ages and that children mainly interact with other children in the same class. We expand care types to include a class setting denoted by *l* and take into account the probability a child is in care type *k* and class *l* based on their age. This is described in further detail in Supplemental Methods.

Based on standards established by large scale contact surveys (6), contact matrices typically count average ‘daily contacts’. Therefore, the proportion of children in age group *i* who receive care of type *k* on any given day can be decomposed into 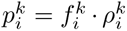, where 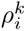 is the proportion of children aged *i* who regularly receive care in *k* (at least once a week), and 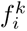 is the average fraction of days of the week they are in care of that type.

### Modeling contacts with caregivers

We also estimate contacts between young children and their caregivers in each type of care. This follows suit from other methods (13) where teachers’ contacts are incorporated as contacts that occur in ‘school’ settings. We follow the same logic as above: contacts are dependent on the probability that a teacher in care setting *k* is of index age *i*, and encounters a contact individual of age *j* on an average day. The formulae for estimating these contacts are summarised in Table S2, and described in further detail in Supplemental Methods.

### Augmenting existing contact matrices

After estimating the average daily contacts between young children, we augment an existing well-cited contact matrix (Mistry et al. (13)) to include these missing childcare contacts. To avoid double-counting, we remove all school-setting contacts for individuals under 5 from this matrix—these are either 0 for those between 0-3 or describe contacts for preschoolers as defined by the American Community Survey (ACS) census—and add our estimated childcare contacts. Note, this procedure reduces the overall number of contacts between 3–4-year-olds and older children relative to the matrix of Mistry et al., which includes these cross-age school contacts, whereas ours does not. The origin of these contacts is unclear, as school interactions between preschoolers and children attending elementary school or higher grades would generally not be expected if contacts were assigned strictly based on the ACS classification.

### Data

To estimate the childcare utilization statistics and characteristics needed to calculate age-specific contacts, we use the 2019 NCES ECPP Study (31) (the most recent version with raw data publicly available). This is a comprehensive National Household Education Survey that estimates early childhood care and education arrangements for children in the United States under the age of five who were not yet enrolled in kindergarten. For each care type respondents are asked a range of questions including the number of hours and days per week on average their children are in care, and the number of other children cared for in each type. The survey questions and responses used in our analysis are provided in Table S5. There were 7092 survey respondents in total.

For children who receive care in formal childcare centers, the ECPP lacks information about other children participating in these settings. Instead, we assumed that children receiving center-based care are arranged into age-based classes. To choose a reasonable, nationally-representative scheme for arranging children into these classes in our model, we reviewed state-specific regulations on minimum caregiver ratios and maximum class sizes by age (reported on state websites and in “Child Care and Development Fund (CCDF) Plan” documents (32)), as well as guidelines for the same from the federal government’s Office of Child Care (33).

To estimate the age and distribution of caregivers, we used summary statistics from the National Survey of Early Care and Education (NSECE) (34, 35), a series of surveys targeted at households, home-based care providers, and classroom staff in early care and education classrooms as a proxy for the age distribution of carers.

### Parameter estimates

We extract the monthly age of the index individual and the proportion of children in age group *i* who routinely receive childcare of type *k*, 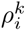 from the ECPP survey.

For each care type *k*, we calculate the average fraction of days per week children spend in these care types, 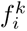. We used a generalised linear model to fit a natural spline with three degrees of freedom to this fraction of days as a function of age, and found that there was no significant trend by age, but the difference between care types was significant (Figure S2). Thus, we used a fixed value for the average fraction of days a child attends each care type, independent of age.

To estimate the average number of children being cared for together in each setting (*n^k,l^*), we use a different approach for home-based and center-based care. For at-home care by a relative or non-relative, we use the mean number of other children present in their child’s care setting based on the ECPP survey. For children in center-based care, we assumed that centers were operating with age-specific groups of children (classes) and that all contacts occurred within these classes. We assumed that the age bands defining each class, as well as the number of students and caregivers per class, were informed by federal regulations on minimum caregiver ratios and maximum class sizes by state-defined age groups. Additional details are provided in Supplemental Methods.

We estimated the conditional age distribution of caregivers given they are in a care setting *k*. For children taken care of at home by a relative, the median age of the caregivers for at-home relative care was 60 years (interquartile range IQR: 52-65), with a range of 11 to 86 (31).

For non-relative home care and center-based care, we used summary statistics from the NSECE (34, 35). Using these data, we fit a smoothing spline and normalise to ensure a probability distribution (see Figure S1). Further, for the probability that an adult in age group *i* routinely works as a childcare provider of any type, we use data from the Bureau of Labor Statistics to obtain the age distribution of persons employed providing “child day care services” (36).

Finally, we approximated the US age distribution by month by taking US Census data (37) and assuming that age is distributed equally over monthly age groups.

### Case study: Measles simulation

We hypothesized that the underestimation of contacts among young children in existing contact matrices would lead to systematic underestimates of the burden of emerging infectious diseases in this age group, and create biases in estimates of the impact of control measures. Motivated by the resurgence of measles in the US and Canada in 2025, we simulated the distribution of outbreak sizes after the introduction of a single infected case of infection into a population of 100,000. We assumed an *R*_0_ value of 12 (38) in the absence of childhood measles-mumps-rubella (MMR) vaccination, and examined two levels (low: 70%, high: 95% coverage) of vaccination coverage for both younger (1-5 years) and older children (6-13 years) leading to four scenarios (Table 1). These values were chosen to represent recent patterns observed in county-level 2-dose MMR vaccination coverage for kindergarteners in the US (39, 40), where low coverage in younger children and higher coverage in older children indicate declining coverage rates (e.g. Portsmouth county, Virginia), and high coverage in younger children and low in older children indicate increasing coverage rates (e.g. Lassen county, California). For all scenarios we assumed 95% vaccination coverage for ages 14-64 since historically national coverage levels have been high in the US (41) and assumed older adults (aged 65+) are fully immune due to exposure and recovery from natural infection. Vaccine efficacy against infection for one dose of MMR was assumed to be 93% and 97% for two doses (42) and was modeled as an all-or-nothing vaccine, that is, a fraction (given by the efficacy) of the vaccinated individuals are fully protected from infection, while the rest remain fully susceptible. Children aged 1-3 years were assumed to have received one dose of MMR and all other age groups had both doses. We assumed an average infectious period of 8 days (43).

**Table 1:** Four scenarios of MMR coverage in children under 13, representing consistently high or low coverage since 2017, or declining or increasing coverage since 2017. Example county-level 2-dose MMR vaccination coverage rates for kindergartners from 2017 to 2023 were extracted from (40).

|  | **High vaccine coverage in older kids (95%)** | **Low vaccine coverage in older kids (70%)** |
| --- | --- | --- |
| **High vaccine coverage in young children (95%)** | Consistently high vaccine coverage (95%) in all children under 13, e.g., Northumberland, PA: average 96% since 2017 | Increasing vaccine coverage since 2017, e.g., Lassen county, CA: 74% in 2017, 96% in 2023 |
| **Low vaccine coverage in young children (70%)** | Declining vaccine coverage since 2017, e.g., Portsmouth county, VA: 97% in 2017 to 70% in 2023 | Consistently low coverage in all children under 13, e.g., Yates county, NY: average 73% since 2017 |

In each scenario we stochastically simulated the spread of measles starting from one infected young child (*≤* 5 years) using age-specific contact matrices produced by Mistry et al. (13), and the augmented version including childcare contacts we present here. For each scenario, we ran 10,000 replicates of the simulation and investigated the distribution of total outbreak size (total measles cases) and distribution of cases by age-groups. Model details are provided in Supplemental Methods.

## Results

### Frequency and characteristics of childcare attendance in the US

On average, 64% of children under 5 were receiving some type of childcare by someone other than their parent or guardian on a regular basis at the time of survey completion; 28% receive home-based care from a relative, 14% are cared for home by a non-relative, and 35% attend a childcare center (Table 2, Figure 2). The proportion of children receiving any type of childcare increased with age - from 40% for infants less than 1 year old to 73% of 4 year olds. For infants, in-home relative care was the most common care type (26% of infants), but for children aged 3-4 center-based care dominated (52%). The proportion of children receiving home-based care increased slightly from birth to around 2 years of age and then decreased for older ages, while the proportion receiving center-based care increased continually (Figure 2). Across all ages, children receiving care spent on average 3.7 days per week in home-based relative care, 3.9 days in home-based non-relative care, and 4.2 days at childcare centers (Table 2, Figure S2A). The average “day” of childcare corresponded to 6.35 hours of care averaging over all care types.

**Figure 2:**
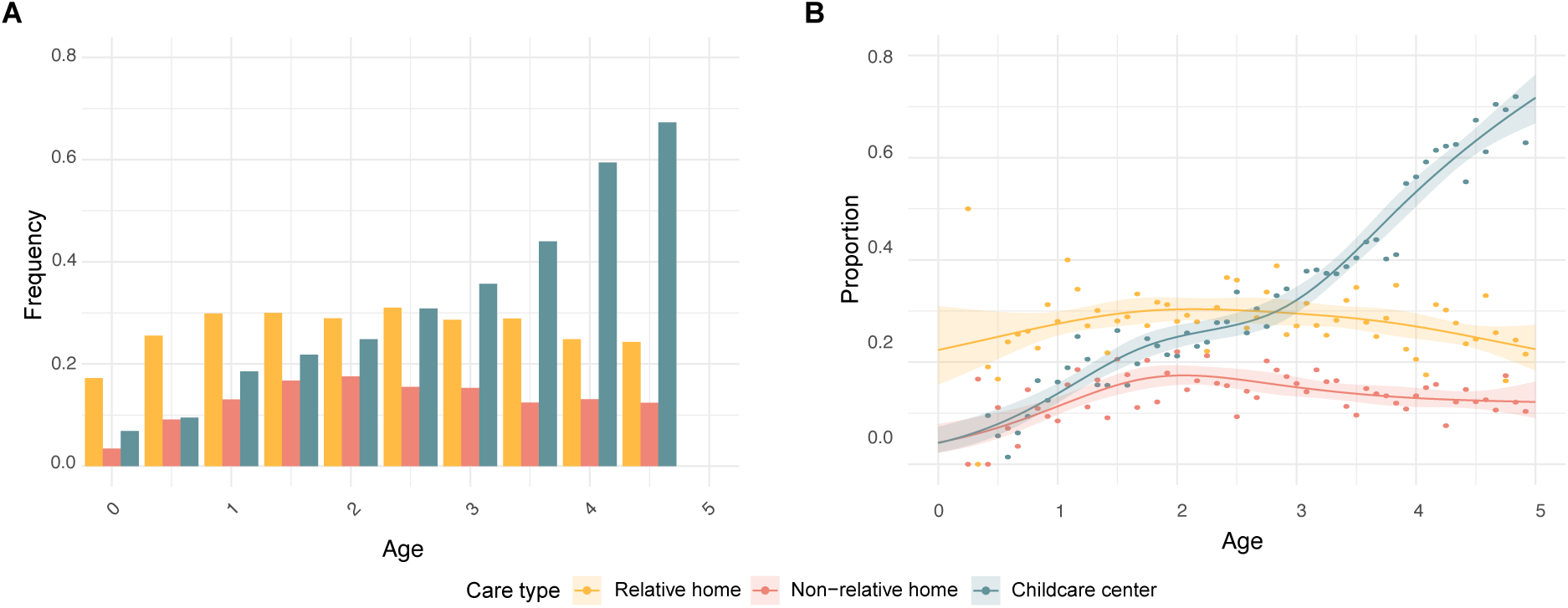
Frequency of childcare attendance in the US by age from ECPP. (**31**) **data.** (A) The frequency of children accessing any type of care (at-home by a relative (yellow), at-home by a non-relative (red), and Childcare center (blue) settings), by six month age bins. (B) Generalised linear model fit (with 4 degrees of freedom) and 95% confidence intervals for the proportion of children accessing each childcare type by age in months.

**Table 2:**
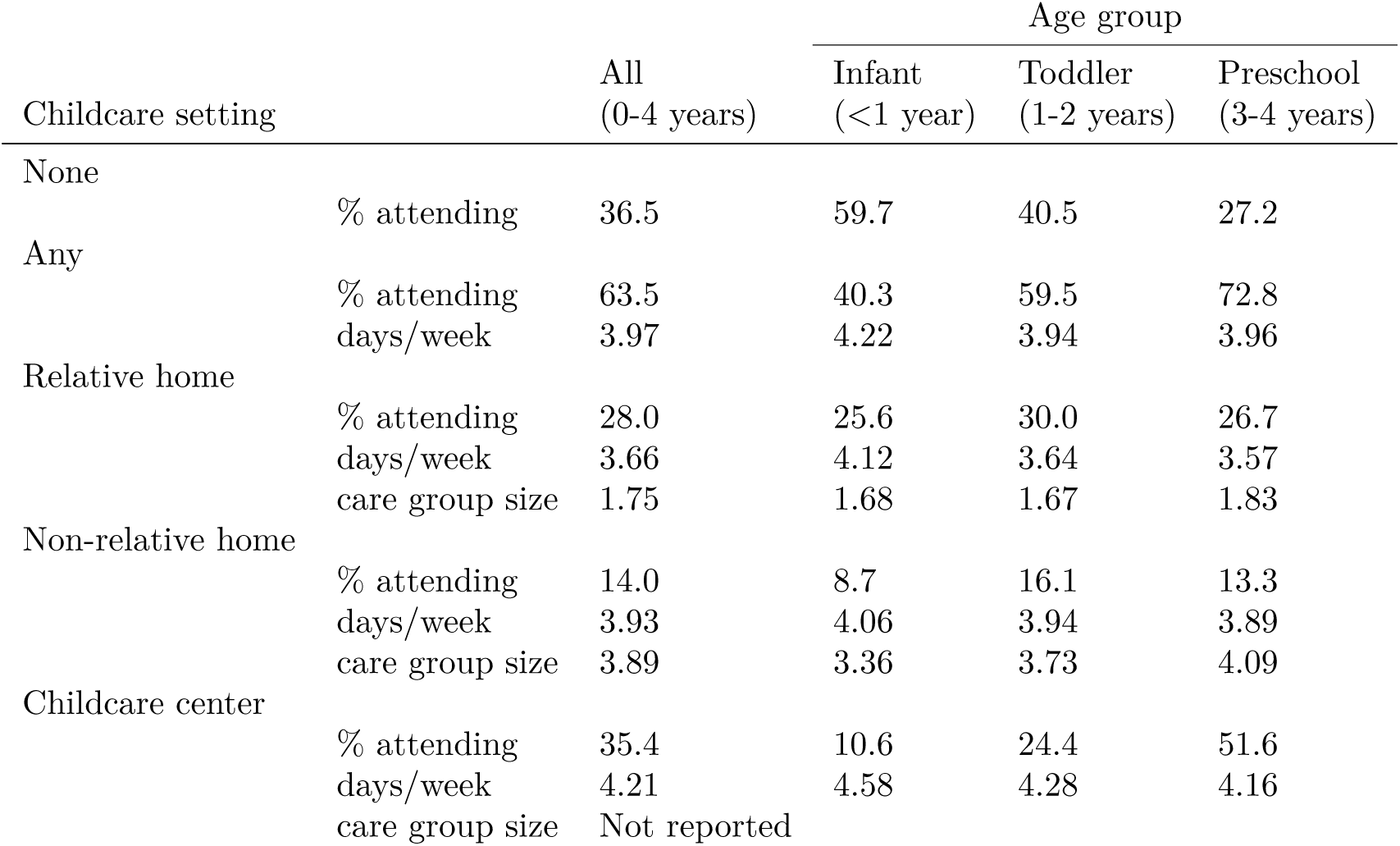
Childcare use summary from ECPP. Ages indicated are inclusive (i.e., 3-4 includes all 3 and 4 year olds).

### Estimated contacts occurring in childcare settings

We used the above data on childcare attendance and characteristics of each childcare setting to construct age-stratified contact matrices in each childcare setting (Figure S3). We estimated that children cared for in-home by relatives contacted an average of 0.8 other children per day compared to 2.7 for children cared for in-home by non-relatives. For children attending childcare centers, estimated contacts varied from 4.6 for infants to 12.9 for 4 year olds. For children receiving home-based care, the contact patterns were determined mainly by the age distribution of children receiving care in this setting, since we assumed home-care settings do not divide children up by age. Children who regularly received home based care from a non-relative had on average 0.3, 0.6, 0.7, 0.6, 0.5 contacts with children ages 0,1,2,3, and 4. For care from relative, contacts were substantially lower, at 0.1, 0.2, 0.2, 0.2 and 0.1 respectively. For center-based care, our inferred contact matrices showed strong associative mixing between ages, due to the division into classes by age (or developmental stage). For example, 2 year olds in childcare centers were inferred to have 3.4 contacts with other 2 year olds, but only 1.4 with 1 year olds (see Supplemental Figure S3).

Combining contacts across settings and taking into account the proportion of US children in each age group who use any (and each) form of childcare, we constructed an overall age-stratified matrix for contacts occurring in childcare settings (Figure 3A). We observed age-based clustering of contacts, increasing for older children, driven by increased prevalence of center-based care as children approach school age. Overall, we estimated that the average daily contacts occurring in childcare settings was 0.4 for infants, 1.1 for 1 year olds, 1.5 for 2 year olds, 2.9 for 3 year olds, and 5.3 for 4 year olds. Infants on average had 0.2 contacts with other infants, 0.07 with 1 year olds, and only 0.04 with 4 year olds. 4 year olds had on average 3.5 contacts with other 4 year olds, indicating the large relative number of contacts in program care. These values are summarised in Figure 3A, and in the associated files in the provided GitHub repository.

**Figure 3:**
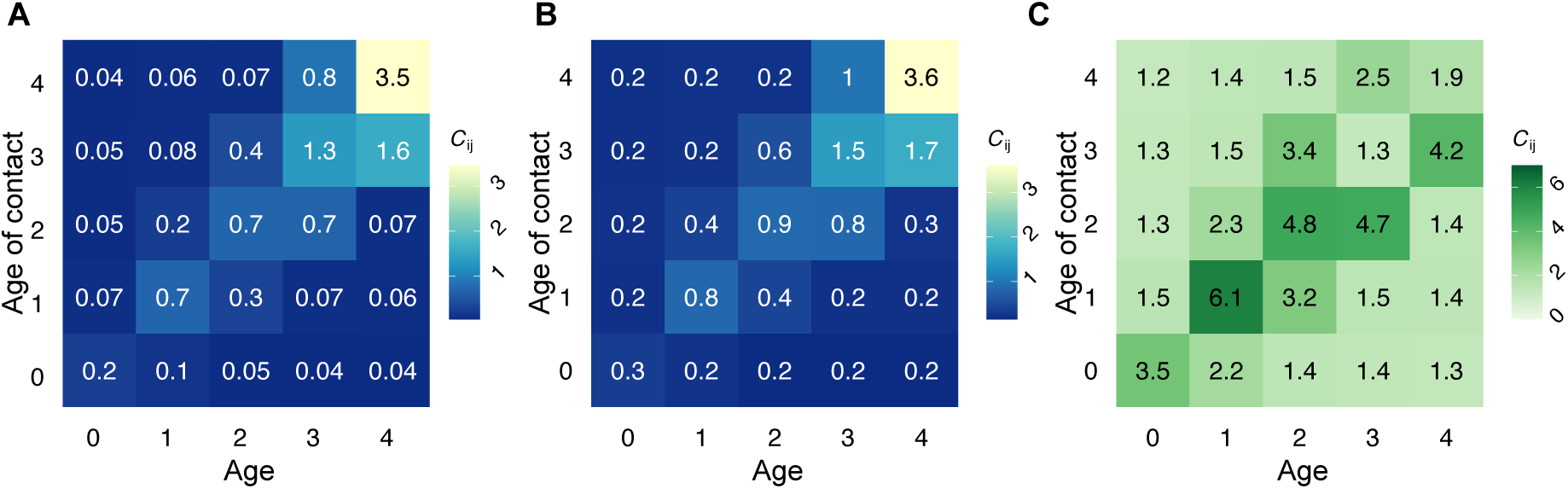
Average daily number of contacts for children under 5 (by yearly age group). (A) Daily number of contacts in childcare settings; and (B) Daily number of contacts in all settings by augmenting existing synthetic matrices in Mistry et al. (13) with childcare matrix. (C) Fold increase in contacts in all settings after augmenting with childcare matrix. The age of the index individual is on the x-axis and the age of the corresponding contact is given on the y-axis.

We augmented the contact matrices estimated by Mistry et al. (13) for young children with our childcare matrix (Figure 3B, see Figure S4 for all age-groups). The total number of contacts in young children increased substantially, with the largest increase seen within age-groups; for e.g., contacts between infants, 1-year olds, and 2-year olds increased 3.5, 6.1 and 4.8 fold respectively (Figure 3C). Overall, contacts among children aged 5 years or younger increased by ∼ 20 - 500%.

### Effect of increased contacts in children in a measles case study

In the measles outbreak simulation we found that across scenarios, the updated matrix with higher contacts among younger children increased the probability and size of outbreaks (Figure 4). Note, since the goal was to compare the effects of the two matrices on measles transmission, we have not included any control measures in the model which would be enacted during real-world outbreaks. The largest difference between the updated with-childcare and the original no-childcare contact matrix occurred under the scenario with low vaccination coverage among younger children and high coverage among older children, where the no-childcare matrix is most likely to substantially underestimate the outbreak size. Differences in age-specific contributions to the epidemic were driven primarily by younger children, whose contribution is increased as expected under a matrix including childcare. For both vaccination coverage levels, the updated matrix—by increasing contacts among young children and reducing between young and older children—results in smaller spillover into older children. The contribution of individuals over 14 years to the epidemic is largely unchanged since the added adult-to-young children contacts in childcare settings don’t affect total adult contacts in a meaningful way (Figure S5). Full distributions of the outbreak sizes broken down by age-groups and mean age-specific attack rates are provided in Supplementary Figures S6, S7.

**Figure 4:**
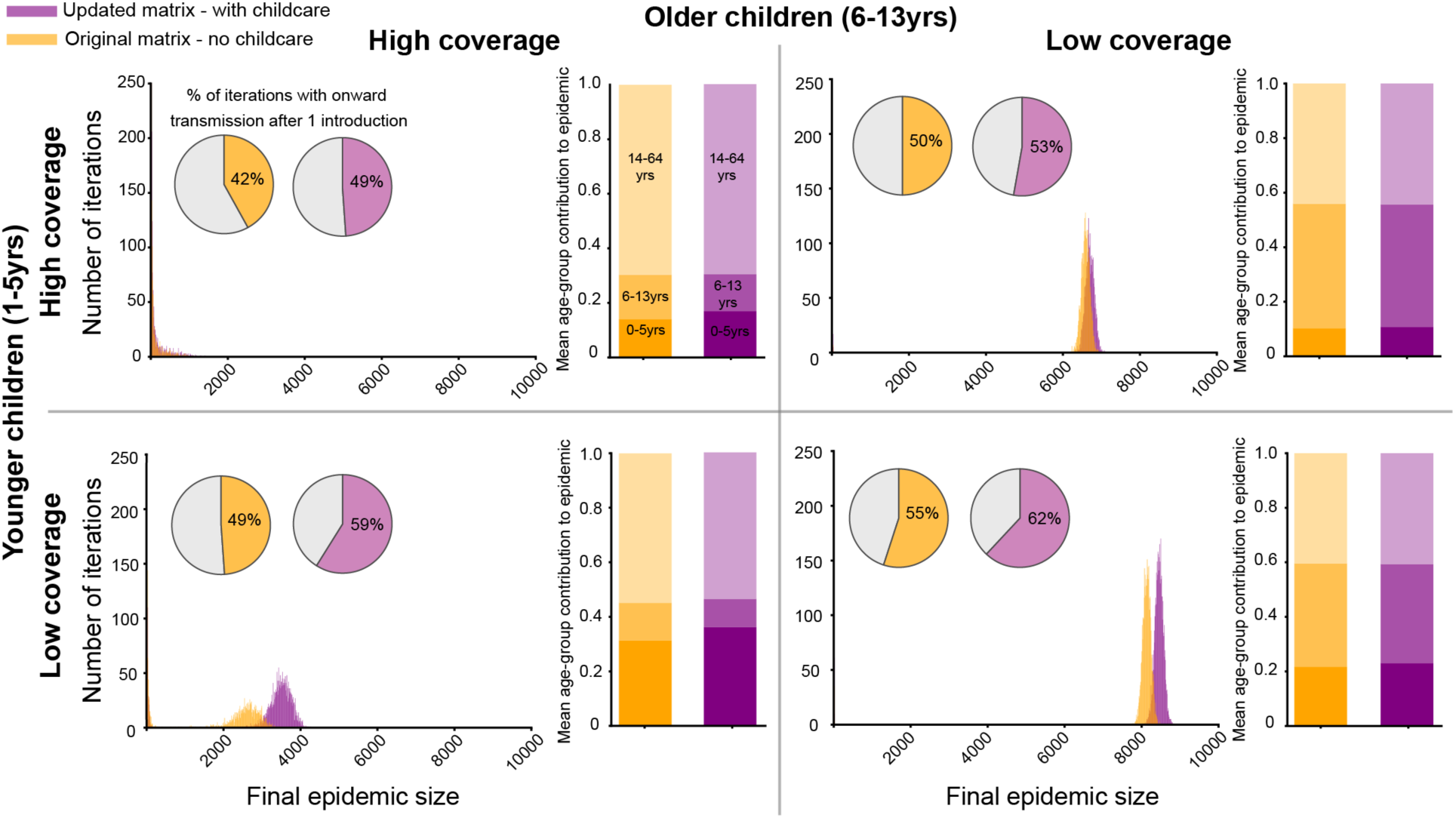
Simulation results of the measles case study across vaccination scenarios. Each of the four panels contains histograms of the final epidemic sizes for iterations where onward transmission occurred after the introduction if a single infection, and bar graphs of the mean contribution per age-group (0-5yrs, 6-13yrs, 14-64yrs) to the total epidemic for both the original - no childcare (orange) and updated - with childcare (purple) contact matrices. Simulation results are for 10,000 replicate introductions each in a population of size 100,000.

## Discussion

In this study we used a national survey of childcare use to update estimates of the frequency of contact among children under age 5 in the United States, and demonstrated the potential impact on disease transmission patterns. Reliable models of age-stratified infection risk and age-targeted interventions require estimates of the frequency of contact between different age groups. However, comprehensive national surveys of disease-relevant contacts have not been conducted in the United States. Existing US contact matrices are inferred from census data - which lacks information on childcare use - and are calibrated using data from contact surveys in European countries (6) - which differ significantly from the US, where there is no federally-mandated paid parental leave, resulting in very early placement of children in non-parental care arrangements. We argue that prior approaches have led to substantial underestimation of contacts among children not yet in formal schooling, a gap we address by creating updated contact matrices that include our estimates of contacts occurring in childcare settings.

We inferred a significantly higher number of average daily contacts between young children compared to values in commonly used synthetic matrices for the United States (13), with sixfold increase among children aged 1 and a 1.2—5-fold increase among all other age groups under age 5. These results align with expected childcare attendance patterns, and studies of levels of infections in young children, especially those in care (44–46). Simulating outbreaks of measles, we found that our updated estimates of contacts among young children substantially increased i) the probability of an outbreak occurring, ii) the overall outbreak size, and iii) the relative contribution of children ages 0-5 vs 6-13, compared to prior estimates. These differences are most pronounced in scenarios with lower vaccination coverage among younger children versus older children, a scenario that may be becoming increasingly common given recent declines in vaccination coverage. These findings highlight the need for accurate representation of contact patterns to inform outbreak planning efforts.

An advantage of the Early Childhood Program Participation survey, compared with census data, is that by reporting children’s age at a monthly resolution, it allows us to estimate contacts at a monthly - vs yearly - age scale. This is especially important for young children, where developmental milestones and fine age scales determine changes in patterns of care (in moving up classes or federally mandated class sizes) and thus exposure to infections. Further, this age scale is important in the context of childhood vaccination policies, where recommendations are defined monthly. For example, a three dose schedule for diphtheria, tetanus, and acellular pertussis is recommended for administration at 2, 4 and 6 months of age, and an RSV monoclonal antibody is seasonally recommended in a child’s first year and thus strongly tied to the monthly age of the infant (47). It is therefore useful to have contact matrices at this finer scale to enable simulation of scenarios for optimizing new and existing interventions for childhood infections.

The results of the ECPP survey and the methods we present here provide important insight into the type of data that should be collected in future purpose-designed contact surveys to reliably estimate average daily contacts in childcare settings. Contact surveys should ask specifically about formal and informal childcare arrangements, with clear definitions of any terms like ‘daycare’ used, and researchers should develop standardized survey methods to ensure coverage of all types of childcare, for all relevant ages. Across the US, there are likely strong geographical differences in contact frequencies for young children, as regulations around childcare group size vary greatly; e.g., Texas and North Carolina allow 20+ 2 year olds per class, Florida allows 40 pre-K students, and Georgia allows 6 infants per caregiver. Conducting contact surveys across different states would help better characterize the different disease dynamics across the country. Globally, any best-practice guidance for conducting new contact surveys should explicitly address how to capture contacts among the youngest age groups, taking location-specific variations in childcare structures into account. Finally, future studies should attempt to directly link contact patterns with epidemiological data to better validate the relationship between contact type, setting, duration, frequency, etc with disease risk.

## Acknowledgments

We thank members of the Johns Hopkins Infectious Disease Dynamics Group and the Insight Net Social Mixing Working group for helpful feedback on this project.

## Data availability

Code for generating the contact matrices and simulations conducted in this study is open source and available on Github: https://github.com/HopkinsIDD/childcare_contacts

## Supplementary Methods

### Interpreting contacts among young children in published contact matrices

By definition, contact matrices used in mathematical models of infectious disease should describe the number, frequency, or duration of contacts that have the possibility of spreading the disease of interest, between two defined groups of individuals. For diseases spread by close spatial or physical contact, it’s usually impossible to measure contacts direction, though some studies have used purpose-deployed physical proximity sensors (1–3) or co-location of mobile devices (4, 5) to approximate contact patterns. More commonly, models used contact matrices derived from “contact diaries”, where individuals record the number of unique individuals with whom they have made contact (of a certain duration, intensity, and physical proximity) in a particular 24-hour period. The most well-known example of this method is the POLYMOD survey covering 8 European countries. No such similarly comprehensive contact survey has been conducted in the US, and so instead, two highly cited papers have attempted to impute a plausible contact matrix by using high-level census data and survey derived contact matrices from other countries (12, 13). However, both studies have limitations with regard to the treatment of contacts between young children.

In Mistry et al (13), the average number of contacts occurring in schools (“school” component of matrix) is populated by combining estimates of the frequency of school attendance with estimates of school sizes. School enrollment is taken from the American Community Survey (the detailed “long-form” census administered to a subset of the population every year) (48). For children ages 3 and older, census respondents can choose “nursery school, preschool” as an option, in addition to “kindergarten”, “grades 1 - 12”, “college undergraduate years” or “graduate or professional school”. For older children, it appears that separate data on size distribution of elementary and high schools (and potentially also the grades they serve, which can vary by region) is used to assign contacts.

There are several major limitations to this approach. The resulting contact matrices have zeros for all contacts between age groups less than 3. It is unlikely – even for individuals aged 3-5 – that all attendance in childcare would be recorded with this answer, as many types of childcare are not colloquially referred to as “preschool or nursery school” in the US. It is unclear how the average number of contacts (assumed equal to total school size) is assigned for students in “preschool or nursery school”. Since students are of course unlikely to contact every single student in their school, total contact number in the school setting are re-scaled using weights the authors estimated. By repeating this procedure for imputing “synthetic” contact matrices for the eight countries that *did* have empirically-derived contact surveys, and then comparing the synthetic matrices to those from the surveys, the authors derived scaling factors for each setting relating the two. However, differences in the way childcare is encoded in the European-based POLYMOD vs the US mean the weights were likely unreliable in a new setting, and they don’t address the issue that children are also organized into age-based classes.

A similar issue occurs in Prem et al (12). The UNESCO data they use specifically “excludes purely family-based arrangements that may be purposeful but are not organized in a ‘programme’ (e.g. informal learning by children from their parents, other relatives or friends is not included)”, which therefore also excludes the most common form of childcare for young children in the US.

Although we focus here on young children, because of their near-absence from existing synthetic matrices, for the reasons mentioned above, “school” contact matrices are likely unreliable for older students too. Students in school spend the majority of time in individual classrooms that are stratified by grade, especially in elementary school, and friendships tend to be highly assortative by age. Confirming this common sense, previous studies that used wearable or mobile-phone based physical proximity sensors in schools found strong evidence of assortativity by class, grade, and age (49). The same was found in detailed contact surveys (children could answer for themselves beginning at age 9 or 13 depending on country in POLYMOD).

The synthetic contact matrices developed in Mistry et al and Prem et al represent an innovative use of existing demographic data and have enabled important mathematical modeling work, and we believe our additional analysis of childcare enrollment data improves these valuable estimates. However, the need persists for more reliable and direct measurements of contacts.

### Modeling contacts in center-based childcare between young children

To calculate the average number of daily contacts for children who receive center-based childcare (i.e. care at a “daycare” or “preschool”), we assume that in most cases children in that care type would be divided up into “classes” based on ages, and that children mainly interact with other children in the same class. In this case, we expand each of these care types to include a setting (such as a class or group) indexed by *l*, and then define 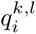 as the *conditional* probability an individual of age *i* who receives childcare of care type *k* is in class/setting *l*. Then, 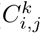 the average number of contacts per day that a child of age *i* has with children of age *j* in care type *k* becomes

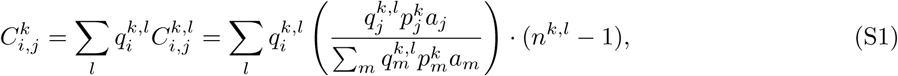

where *n^k,l^* is now the average number of children per class setting *l* in childcare type *k*. For example, imagine we are interested in estimating the contacts for children 0-11 months old in childcare centers, and assume that all centers have infant classes with 6 children total (*n*_childcare center, infant class_ = 6) and 100% of children under age 1 are in this class 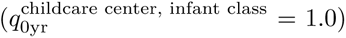. If we assume that childcare center rules are such that children are eligible to move up to the toddler class any time after they reach age 12 months, but they may not move immediately on their birthday due to space constraints or developmental milestones (e.g., not yet walking), then our calculation would additionally need to use - for example - the fact that 10% of children in the 1 year old age group are still in the infant class 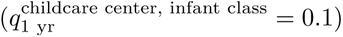, along with the overall distribution of age groups within childcare center settings 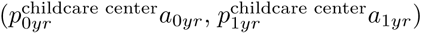. The specific scheme we use for assigning age-based classes, and the data that informs it, is described in the subsequent ‘Parameter Estimates’ section.

### Modeling contacts with caregivers

In addition to the child-to-child contacts, we also include contacts between these young children and their caregivers in each type of care. This follows suit from calculations undertaken by others (13) where teachers were removed from the working population and their contacts made at work were instead incorporated into the school setting matrices. First, for care types where all individuals are in a single group (e.g., at-home care where children are *not* separated into classes), we assume that that there is only a single caregiver. That is, there are no adult-to-adult contacts (note that this can be relaxed). In these care types *k*, if the index individual is a young child of age *i*, the number of contacts with a caregiver of age *j* in setting *k* is the probability that a caregiver in care-type *k* is in age group *j* multiplied by number of caregivers in that setting, that is

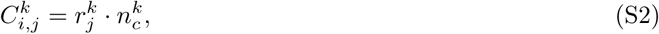

where 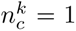 is the number of caregivers in these specific settings of care-type *k* and 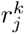 is the probability that a caregiver in care type *k* is of age *j*.

Conversely, if the index individual is a caregiver of age *i* and the contact is a young child of age *j*, this becomes the age distribution of children in this care type of a given size

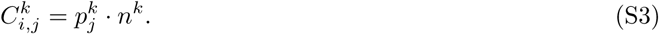

Second, for center-based childcare where children are likely divided into classes, the number of contacts between young children of age *i* with adult contacts of age *j* is dependent on the the conditional probability that the child is in that class, the probability that the caregiver is of age *j* and the number of caregivers in that class *l*. This assumes that children interact with each caregiver in their class equally. This gives

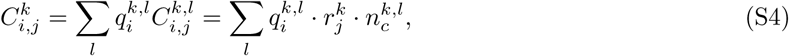

where 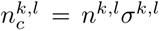 is the number of caregivers per class, decomposed into the number of children per class and the caregiver:student ratio *σ^k,l^*. For the corollary contacts of adult caregivers of age *i* with young children of age *j* this becomes,

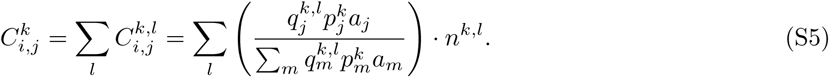

We also here model contacts between caregivers. We assume that caregivers in at-home care by a relative or non-relative are sole-carers and there are therefore no contacts between adult caregivers. For adults who work in childcare centers, we assume for simplicity that caregivers only contacts others in the same class (given a lack of data regarding overall ‘school’ size of childcare centers). Then, the number of contacts between caregivers in class *l* is given by

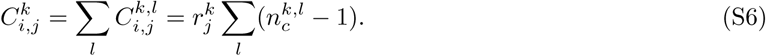

The formulae used to construct these contact matrix entries are summarized in Table S2.

### Parameter estimates

This section is a more detailed version of the ‘Parameter estimates’ section from the main text.

To impute age-specific contact matrices from ECPP data, we need the age of the respondent’s child at the time the survey was completed. The publicly-available data includes month and year of birth, but no the survey completion date. We estimated age using the reported birth month and year, assuming every survey was completed in August 2019. In reality, survey responses were returned anywhere between January and August 2019. This method likely overestimates ages, on average, but avoids having any negative ages. Note that individuals with children up to age 6 by Dec 2018 who were not yet in kindergarten were asked to participate. However, we focus on individuals age 0-4 in this analysis, since these are children most likely to be in care (vs formal schooling) and since they represent 95% of the data

We constructed contact matrices using monthly age group bins, in contrast to prior work that reported only yearly age groups. For disease applications where there are sharp age-specific differences in immunity (due to, for example, infant vaccination schedules), narrower age bands are commonly used in models and childcare is likely the setting where contacts vary most dramatically with age for younger children. For modeling applications where wider age groups can be used, the resulting matrices can be aggregated. Although the survey includes children up to age 7, we focus on individuals age 0-4 in this analysis, since these are children most likely to be in care (vs formal schooling) and since they represent 95% of the data. Note that individuals with children up to age 6 who were not yet in kindergarten were asked to participate in early 2019, so children could have been up to age 7 by the time the survey was returned)

To determine the proportion of children in age group *i* who routinely receive childcare of type *k*, *ω^k^*, we calculate the frequency of responses regarding program attendance in the ECPP. The survey question is a binary response to whether children are receiving care at each of the following childcare types: at home from a relative (“relative home”), at home from a non-relative (“non-relative home”), or attending a daycare center, preschool or pre-kindergarten not in a private home (“childcare center”). The proportion is calculated as the fraction of ‘yes’ responses over the total respondents, as a function of age *i*. Since children may receive childcare in more than one setting, these responses may sum to more than 1 for any particular age *i*.

For each of these care types, respondents are asked how many days and hours each week the child receives care in this care type. We use the responses to this question to calculate the average fraction of days per week children spend in these care types, 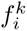. We used a generalised linear model to fit a natural spline with three degrees of freedom to this fraction of days as a function of age (in monthly bins), and found that there was no significant trend by age, but the difference between care types was significant (Figure S2). Thus, we used a fixed value for the average fraction of days a child attends each care type, independent of age (Table S3). Although we found vidence of some bimodality in hours of care per day of care (Figure S2C), suggestive of half-day care for some children, for consistency with contact surveys which count daily contacts regardless of their duration over a threshold (e.g. > 15 minutes) we ignored this in our calculations.

To estimate the average number of children being cared for together in each setting (*n^k,l^*), which determines an index child’s total contacts with other young children while in that care setting, we use a different approach for home-based care and center-based care. For at-home care by a relative or non-relative, respondents in the ECPP were asked how many *other* children were present in their child’s care setting. Survey responses were categorized into [None, 1-2, 3-5, 6 or more]. To obtain the average of the responses, we assume these responses correspond to average class sizes of [1, 2.5, 5, 7].

For children in center-based care, we assumed that centers were operating with age-specific groups of children (classes) and that all contacts occurred within these classes. We assumed that the age bands defining each class, as well as the number of students and caregivers per class, were informed by the state-specific regulations on minimum caregiver ratios and maximum class sizes by state-defined age groups ((32)) as well as federal recommendations (33).

Although there was substantial variation across states, we were able to choose a representative center structure that we believe represents well the approximate situation in centers across the country. We divided centers into fives classes: *l* ∈ *{* 0-11 months (infant), 1 years old (younger toddler), 2 years old (older toddler), 3 years old (preschool), 4+ years old (pre-kindergarten) *}* (Table S4). For the infant and toddler classes, we assumed that children could move up classes after their birthdays but not before, and that there was some variation in the exact age they advance classes (e.g. due to space constraints at centers, use of developmental milestones like walking or potty training for advancement, and parental preferences), so that 25% move up during their first month in the new age group, another 25% in the second month, another 25% in the third month, and the final 25% in the fourth month. For the preschool and pre-kindergarten classes, we assume that centers switch to moving children up as a cohort based on their age by a defined start of the school year (e.g. Sept 1), so that the exact age at which children move up classes is evenly distributed across the months of that age group. In reality, the cut-off between age groups varies somewhat by state (for example, NY has separate ratios for infants <6 weeks; in IL, the infant designation continues until 15 months; in CA all children 2 and over are governed by the same rules). Within each age group, we chose to use the federal guidelines for the class sizes and caregiver-to-student ratios: 6 children per class for infants, 8 per class for toddlers (younger and older), 14 for preschoolers, and 16 for pre-kindergarten, with 2 teachers per class in each age group (Table S4). While some state-specific regulations allow much higher class sizes and caregiver:student ratios (e.g., Texas and North Carolina allow 20+ 2 year olds per class, Florida allows 40 pre-K students, and Georgia allows 6 infants per caregiver), we use the federal guidelines to provide a minimum bound for the number of contacts young children may have on average across the United States.

We next estimated the conditional age distribution of caregivers given they are in a care setting *k*, 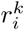. For children taken care of at home by a relative, ECPP provided the age of the caregiver (in years). The median age of the caregivers for at-home relative care was 60 years (interquartile range IQR: 52-65), with a range of 11 to 86. However, ECPP does not ask the specific age of at-home non-relative caregivers, instead providing a binary >18 or under 18 option: 98.9% are *↗* 18. For center-based care, no information on caregiver age is collected in the ECPP. Thus for both non-relative home care and center-based care we estimated the age distribution of caregivers using the NSECE. From this, we used summary statistics from the NSECE (34, 35), where age is categorized for unlisted and listed home-care, and center-based care settings. We took the mean of unlisted and listed home-care as a representative value for our non-relative care setting, and the centre-based care setting. Using these data, we fit a smoothing spline (see Figure S1). Further, for the probability that an adult in age group *i* routinely works as a childcare provider of any type, we use data from the Bureau of Labor Statistics to obtain the age distribution of persons employed providing “child day care services”.

We approximated the US age distribution by month by taking US Census data (37) and assuming that age is distributed equally over monthly age groups.

### Additional assumptions of contact model

There are several additional assumptions and limitations to our approach to translating data from the ECPP survey into age-specific contact matrices for young children.

Based on standards established by large scale contact surveys (6) - which asked index individuals to name contacts they encountered in the prior day that met a particular threshold level of interaction - contact matrices typically count average ‘daily contacts’. Thus, if two individuals are in contact only 3 out of 7 days per week, this counts as 0.43 daily contacts. Most children only attend childcare arrangements a subset of days of the week, which we assume reduces not only their contacts, but also the total number of other children they may encounter when with their caregiver. In essence, this is equivalent to assuming that children are equally likely to be attending childcare any day of the week, which has limitations. It’s likely that the majority of children attend childcare only on the five weekdays, not weekends, and thus their days of attendance are correlated with those of other children, increasing their likelihood of receiving care on the same day and thus being in contact. Moreover, for licensed childcare centers and perhaps also some home caregivers, children accessing part-time care are enrolled in an organized way such that the center always has their maximum number of children. For example, if one child only comes Monday, Wednesday, and Friday, they may accept another child that only comes Tuesdays and Thursdays.

For children who use multiple types of care in a typical week (e.g., 1 day of childcare center and 1 day with a family member), we sum these days to get the total days. However, we may be overestimating total days in care, as an alternative scenario is that both types occur on the same day (i.e., a family member picks up a child from a childcare center to provide after-hours care at their home).

For home-based childcare, respondents in the ECPP were asked how many other children were present in their child’s care setting. Survey responses were categorized into [None, 1-2, 3-5, 6 or more]. To obtain the average of the responses, we assume these responses correspond to average class sizes of [1, 2.5, 5, 7], which is imprecise. We could have alternatively estimated a parametric uni-modal distribution of class sizes and fit it to this data, but in the absence of more data went with the simplest approach.

In the creation of contact matrices for children in center-based childcare, we assumed all interactions occurred within the class structure we created. However, children may also be able to “contact” children from other classes - either by direct interactions in common areas like hallways, playgrounds, etc, or indirect interactions like shared airflow or use of contaminated objects. In addition, we have not taken into account the tremendous regional variation in regulations around the class size in childcare centers.

For home-based childcare, we have assumed that there is always a single caregiver for the entire group of children accessing care in that location, though in reality there could be multiple; for example, with cohabiting family members sharing in caregiving or with a home daycare center that has hired additional employees.

Finally, an unfortunate aspect of the design of the ECPP survey is that respondents did not actually report the exact age of the child at the time the survey was completed on their behalf. Instead, they reported birth month and year, but their individual completion date is not shared with any of the publicly-available data. For the analysis reported here, we assumed every survey was completed in August 2019 (in reality, the survey was conducted between January and August 2019), the last possible date. This means our estimates of age are likely slightly overestimated, and that our estimates of total contacts for a given age are likely underestimated, since contacts tend to increase with age. This will have a larger effect on the assumed monthly age (and thus monthly contacts) of children less than one year of age. This is a major limitation of the current data. While survey receipt date is recorded in restricted-use datafiles, there is currently no way for researchers to request access to those.

Our method for calculating the average number of contacts between age groups in each childcare setting (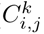, (S3)) assumes that, on average, the index child *i* contacts others *j* according to the full age distribution of children in that care setting, which includes the index child themselves. This is a slight simplification, because in reality the age distribution of contacts should exclude the index individual. If each care setting has a small number of children and if children tend to be non-randomly sharing caregivers (e.g. if same age children vs age-spaced children are more likely that random to share a caregiver), then this assumption could be more important. However, properly accounting for this would require a much more complicated calculation in which the age distribution of contacts is calculated conditional on both index individual age and total number of children in the care setting, and the dataset is likely not large enough to expand the calculation in this manner and maintain reasonable numbers of observations.

To facilitate use in mathematical models, we combine our estimates for contacts in childcare settings with pre-existing contact matrices estimated for other settings (including household) (Figure S4). However, its possible we may be double-counting contacts among some siblings if they attend childcare together, as ECPP does specifically ask about whether the other children who share the care-giver are siblings or non-siblings. Future surveys designed for the purposes of quantifying total infection-relevant contacts should make sure to count only non-sibling peers in care.

Finally, in the analysis of ECPP conducted internally by the National Center for Education Statistics (NCES), individual data points are weighted in an attempt to account for biases in responses to the survey. We did not employ those weights in our analysis. Overall, NCES found a slight bias towards responses by individuals identifying as white, more highly educated, and with children over 1 year of age, though the absolute effect was small (see Ch. 7, 8, (50)).

### Measles simulation model

We use a density-dependent age-stratified SIR model for the measles simulations,

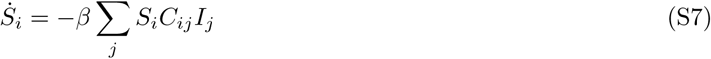

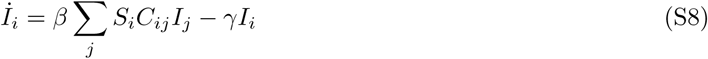

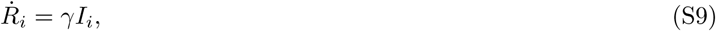

where, *i* = 1*, …, n* are the number of age-groups, and *S_i_*, *I_i_* and *R_i_* are the number of susceptible, infected and recovered individuals per-age group in the population. We assume the population size per-age group *N_i_* = *S_i_* + *I_i_* + *R_i_* is fixed and the total population is *N* = *N_i_*. *C* is the age-mixing contact matrix with the element *C_ij_* corresponding to the daily number of contacts between an individual of age *i* with an individual of age *j*. For simplicity we assume that the infectivity *β* (probability of transmission per infectious contact per day) and rate of recovery *γ* (probability per day an infected individual recovers) are not age-dependent. Using the next-generation matrix method (51), the basic reproduction number *R*_0_ for the model is the leading eigenvalue of the matrix,

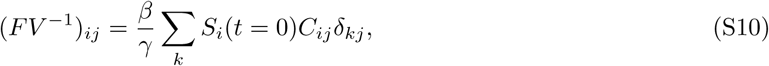

where *δ* is the Kronecker delta function. We fix *R*_0_ and *γ*, and calculate *β* using Eq. S10. Note, this model can be written in a fully vectorized form as,

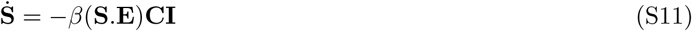

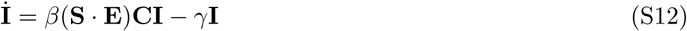

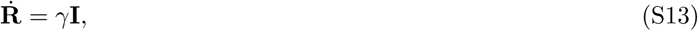

where, **S**, **I**, **R** are (*n* x 1) vectors, **C** is the *n* x *n* contact matrix, and **E** is a (*n* x 1) vector where each entry is one of the basis matrices of the identity matrix; for example if *n* = 2,

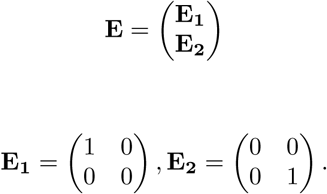

Re-writing the model in this form makes it easy to generalize it for any number of age-groups. We implement a stochastic version of this model using the Tau-Gillespie algorithm (52) in Python using JAX-NumPy (53) to optimize the speed of the simulations.

### Links to Early Childhood Participation Program survey 2019

- [Questionnaire](https://nces.ed.gov/nhes/pdf/early/2019_ecpp.pdf)
- [Reports on data](https://nces.ed.gov/use-work/resource-library/report/first-look-ed-tab/early-childhood-program-participation-2019)
- [Data File Users’ Manual](https://nces.ed.gov/pubs2021/2021030rev.pdf)
- [Public-Use Data](https://nces.ed.gov/datalab/powerstats/151-early-childhood-program-participation-2019) (login required)
- [Restricted-Use Data](https://www.researchdatagov.org/product/5944)

### Supplementary Figures

**Figure S1:**
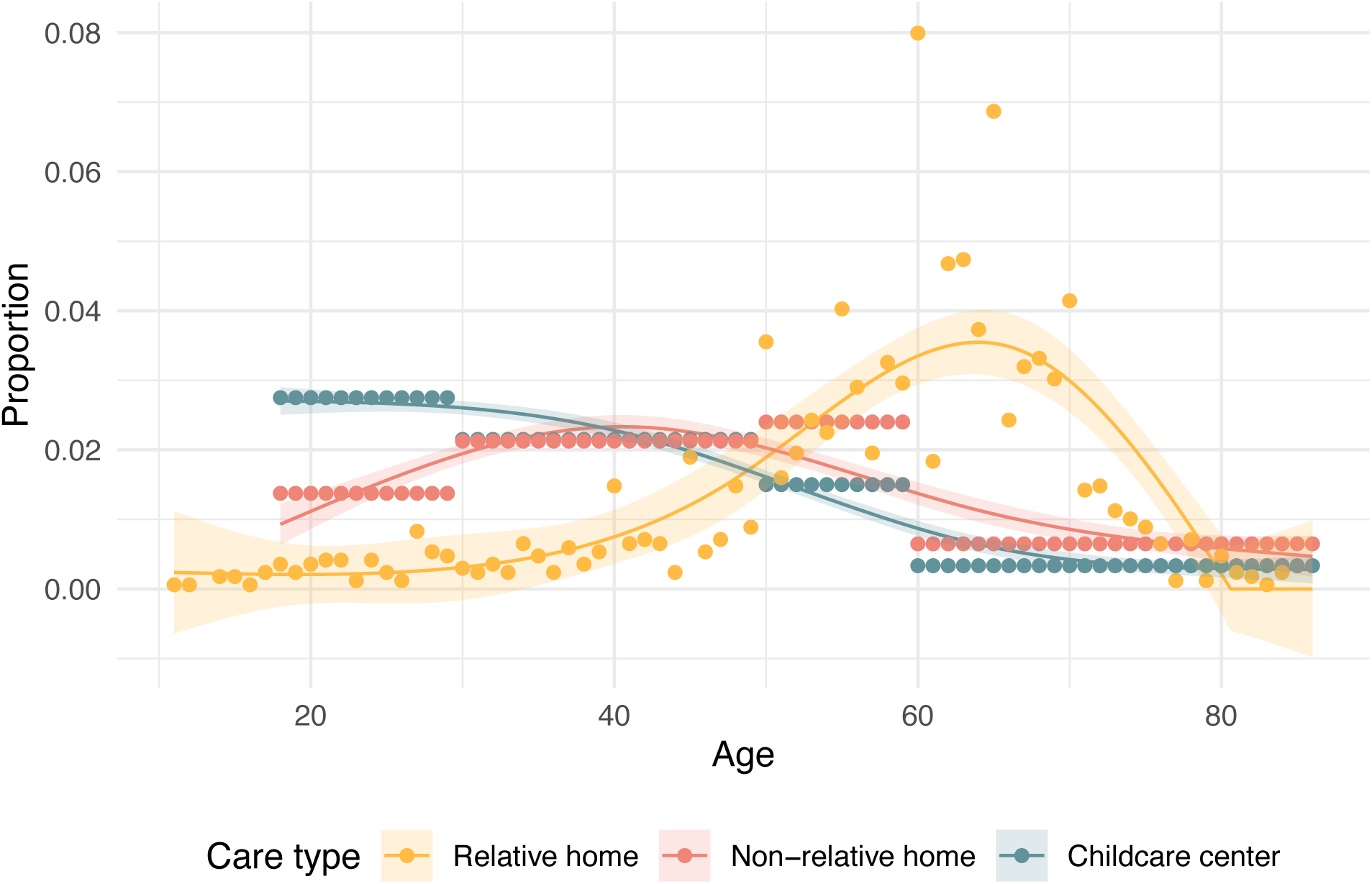
Age distribution of caregivers of young children. The age distribution of caregivers in each care type. The age of at-home relative caregivers is taken from the ECPP (31), and that of at-home non-relative caregivers and center-based caregivers are taken from the NSECE (34, 35).

**Figure S2:**
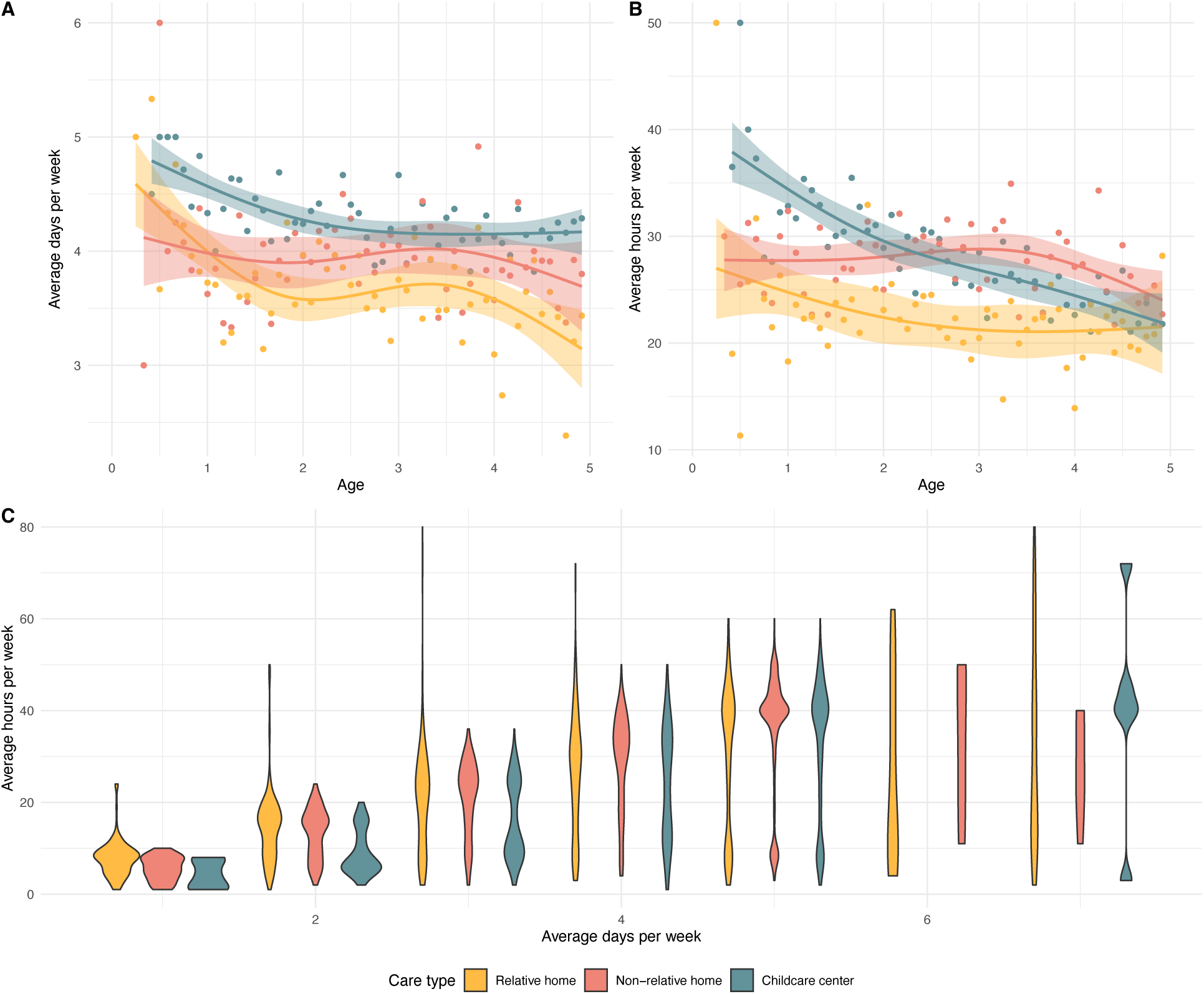
Time spent by young children in different childcare settings. From Early Childcare Participation Program survey, the (A) average days per week, and (B) average hours per week by age, with a generalised linear model fit with three degrees of freedom for each care type. (C) The average hours per week in care as a function of the average days per week, showing bimodal distribution for some days, indicating half-days of care.

**Figure S3:**
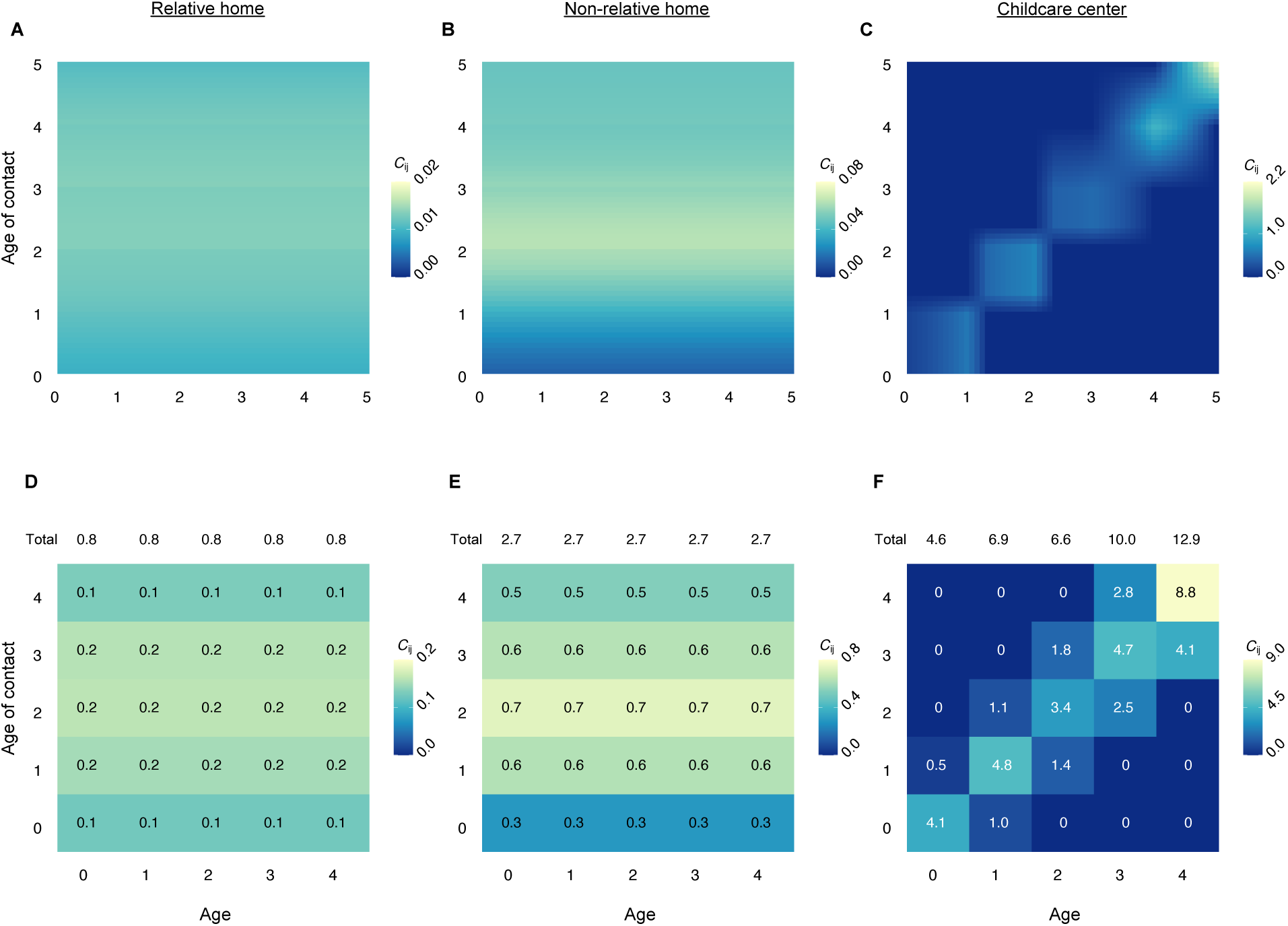
Setting-specific contact matrix for young children in childcare. The average daily number of contacts for index individual of age *i* (columns) with contact of age *j* (rows), for (A, D) In-home care by a relative, (B, E) In-home care by a non-relative, and (C, F) Childcare center. The first row of panels (A-C) are setting-specific contact matrices at a monthly age-scale, and the second row of panels (D-F) show the corresponding yearly matrices, with the total contacts each index individual has with all others under 5 in that setting shown in the top row of each panel.

**Figure S4:**
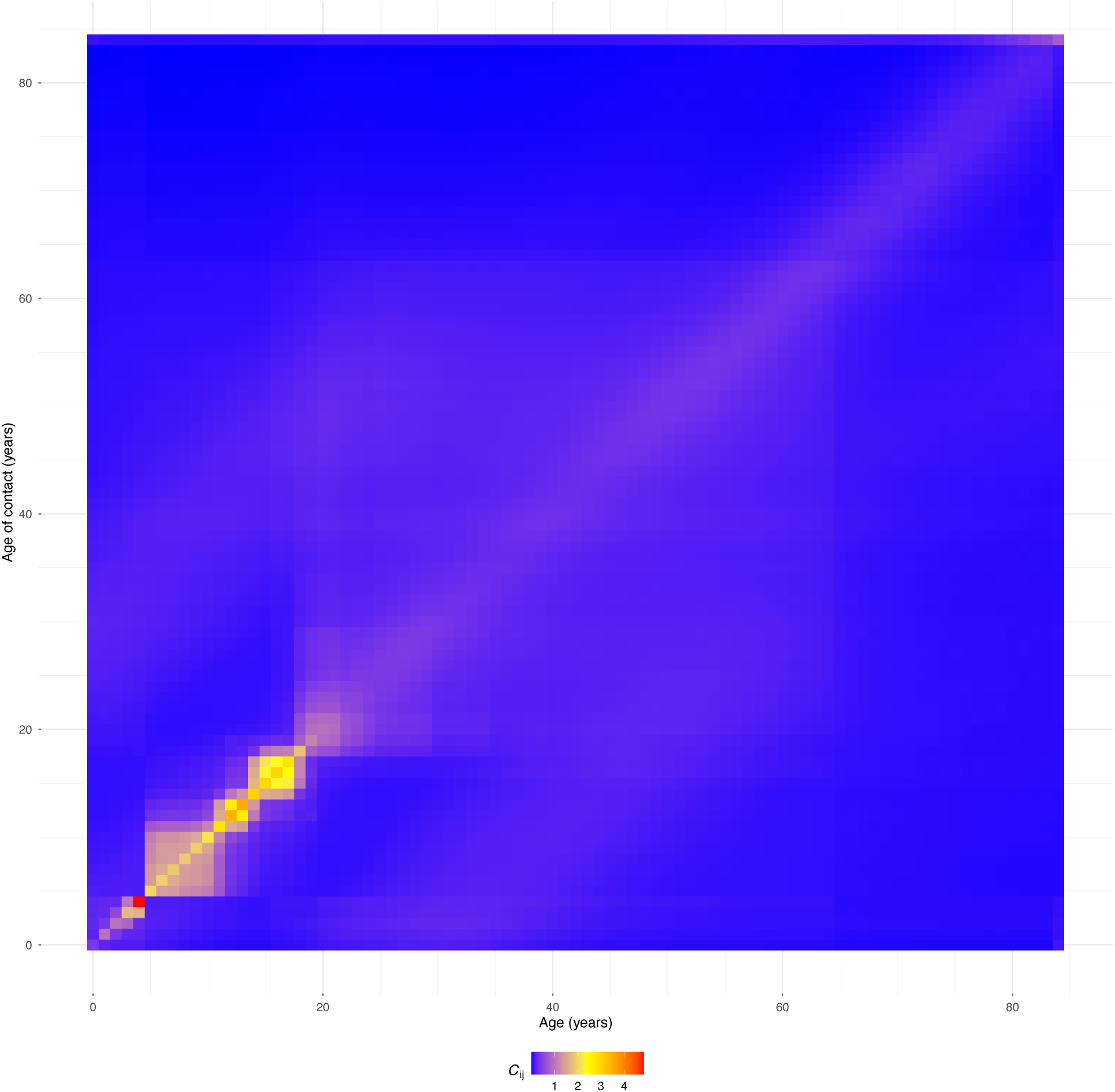
All-age all-setting contact matrix augmented with childcare contacts. The all-setting contact matrix for the US presented by Mistry et al. (13), which includes imputed contacts occurring at home, work, school, and community, was augmented by first removing “school” setting contacts for all individuals under 5, and then adding the childcare setting contacts estimated in this paper. This augmentation includes contacts occurring between children aged 0-4 and childcare providers.

**Figure S5:**
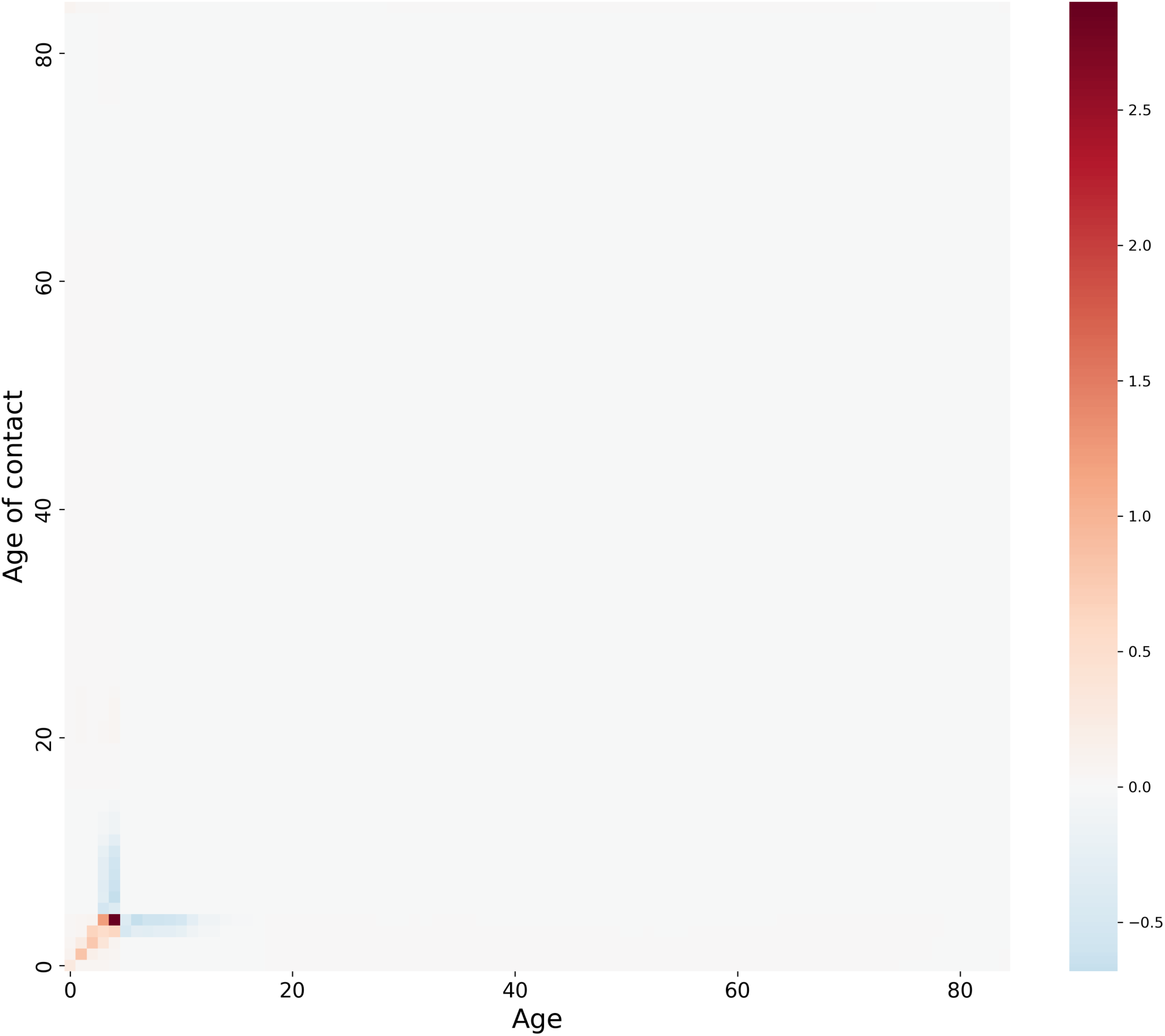
Difference between the augmented all-age all-setting contact matrix with the one presented in Mistry et al. Augmented contact matrix minus the one presented in Mistry et al. Red and blue cells correspond to increase and decrease in contacts in the augmented matrix compared to Mistry et al. respectively.

**Figure S6:**
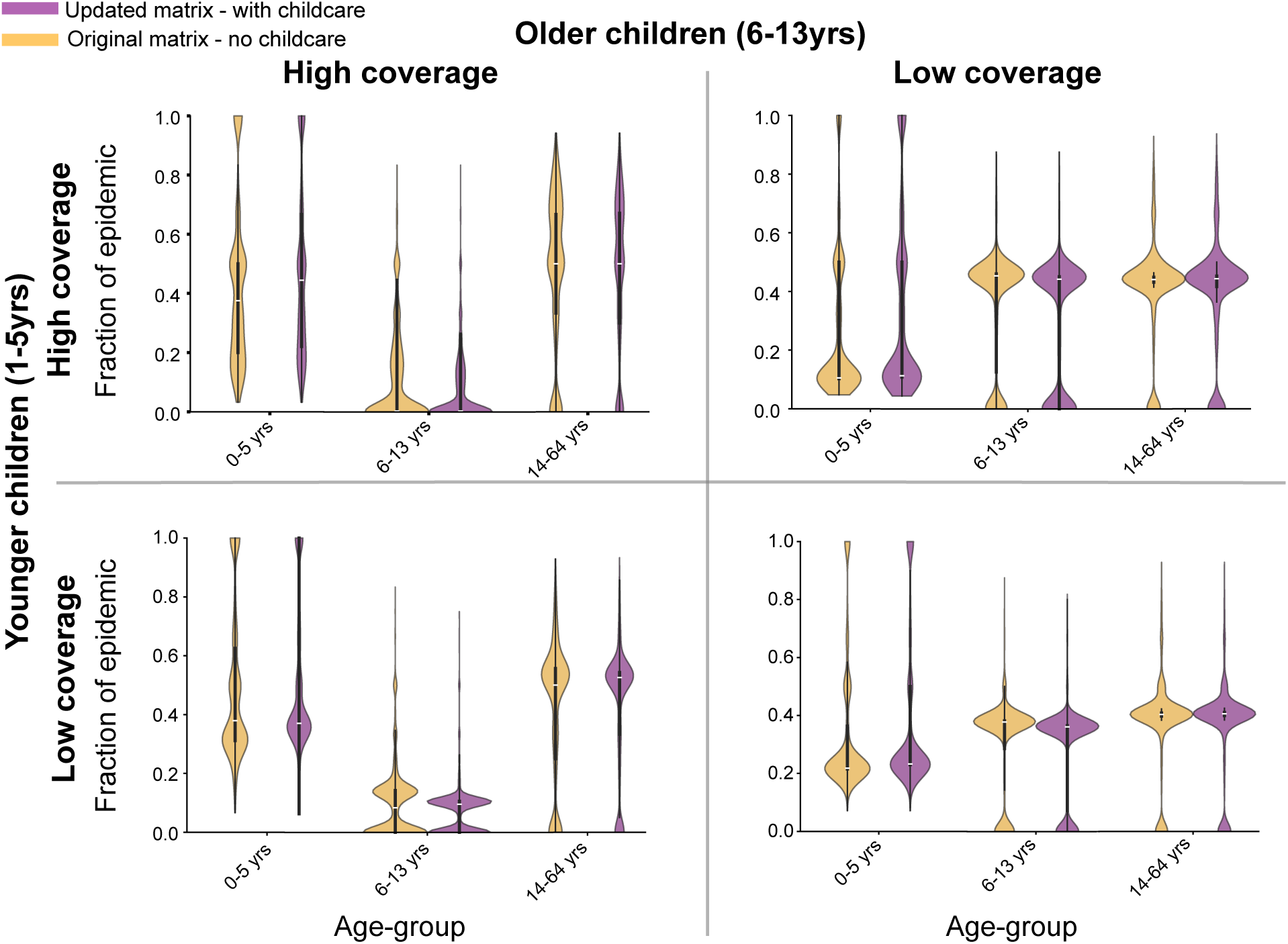
Age-group contribution towards the epidemic for the measles simulation case study across vaccination scenarios. Each of the four panels contains violin plots of the fraction of the total epidemic size belonging to each age-group (0-5yrs, 6-13yrs, 14-64yrs) for iterations where onward transmission occurred after 1 infection was introduced for both the original Mistry et al (orange) and updated (purple) contact matrices. Simulation results are for 10,000 iterations in a population of size 100,000.

**Figure S7:**
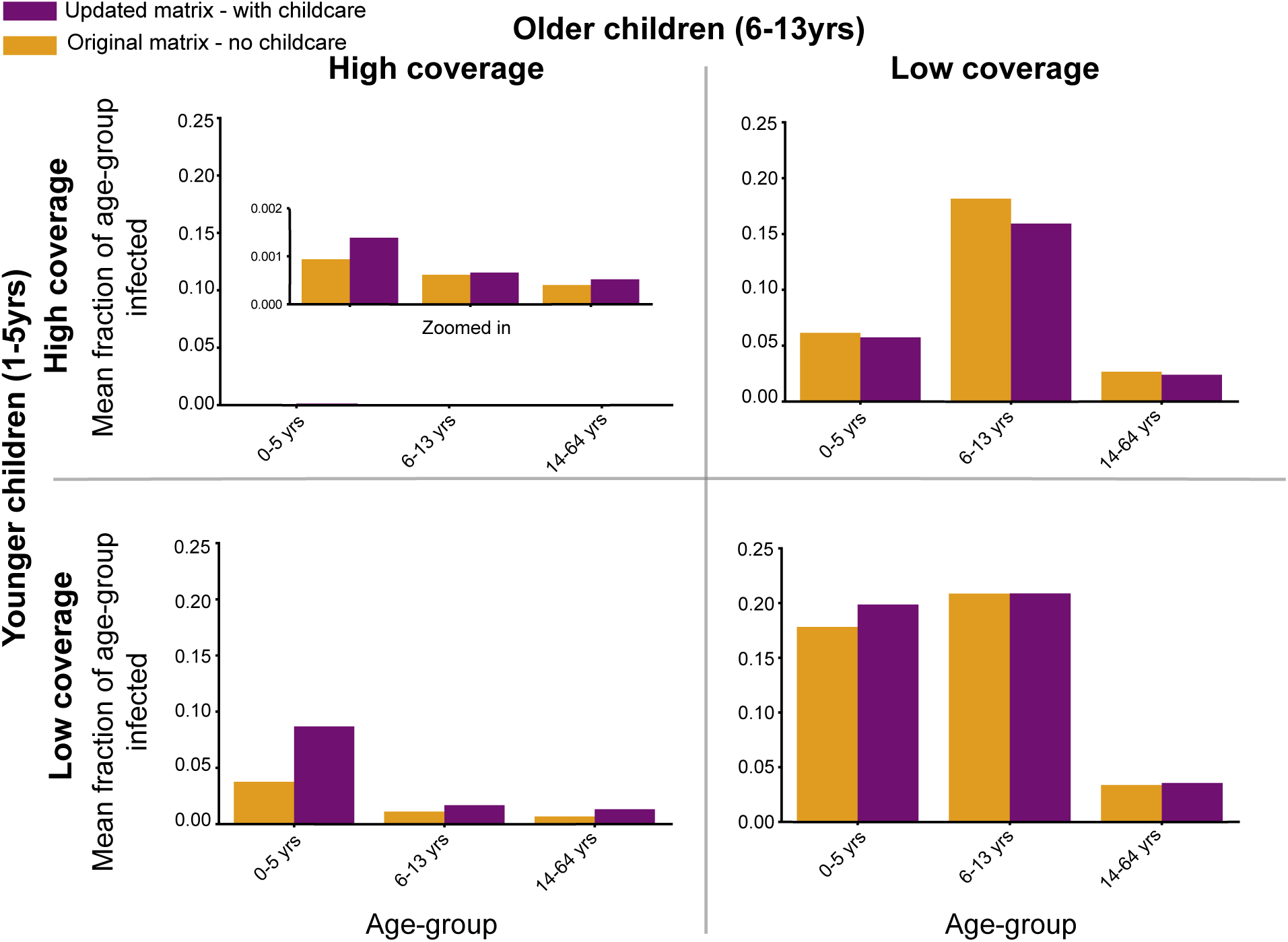
Mean age specific attack rates in the measles simulation case study across vaccination scenarios. Each of the four panels contains bar graphs whose height is the mean of the fraction of the age-groups (0-5yrs, 6-13yrs, 14-64yrs) infected for iterations where onward transmission occurred after 1 infection was introduced for both the original Mistry et al (orange) and updated (purple) contact matrices. Simulation results are for 10,000 iterations in a population of size 100,000.

### Supplementary Tables

**Table S1:** Parameter definitions, units and data sources for calculating average number of contacts on a given day of care.

| Parameter | Description | Units | Source |
| --- | --- | --- | --- |
| $C_{i,j}$ | Average contacts that each child of age $i$ has with children of age $j$ | # of contacts per day | Calculated |
| $C_{i,j}^k$ | Average contacts that each child of age $i$ has with children of age $j$ in childcare type $k$ | # of contacts per day | Calculated |
| $p_i^k$ | Probability child in age group $i$ routinely receives childcare of type $k$ on any given day | [0,1] | Early Childhood Program Participation (ECPP) Survey 2019 (31) |
| $f_i^k$ | Average fraction of days per week child of age $i$ spends in childcare type $k$ | [0,1] | Early Childhood Program Participation (ECPP) Survey 2019 (31) |
| $\rho_i^k$ | Proportion of children in age group $i$ who receive childcare of type $k$ regularly | [0,1] | Early Childhood Program Participation (ECPP) Survey 2019 (31) |
| $n^k$ | Average number of children in childcare type $k$ | # kids | Childcare.gov Supervision: Ratios and Group Sizes (Recommendations), 2025 (33) |
| $q_i^{k,l}$ | Conditional probability child in age group $i$ who receives childcare of type $k$ is in class $l$ | [0,1] | |
| $a_i$ | Fraction of the entire population in age group $i$ | [0,1] | United States Census Bureau, National Population by Characteristics, 2025 (54) |
| $n^{k,l}$ | Average number of children per class $l$ in childcare type $k$ | # kids | Childcare.gov Supervision: Ratios and Group Sizes (Recommendations), 2025 (33) |
| $\sigma^{k,l}$ | Caregiver:Student ratio in class $l$ in care type $k$ | # adults / # kids | Childcare.gov Supervision: Ratios and Group Sizes (Recommendations), 2025 |
| $r_i^k$ | Probability that an adult who works as carer of type $k$ is age $i$ | [0,1] | National Survey of Early Care and Education (NSECE) 2019 (34, 35) |
| $n_c^k$ | Average number of caregivers per childcare type $k$ | # adults | Childcare.gov Supervision: Ratios and Group Sizes (Recommendations), 2025 (33) |
| $n_c^{k,l}$ | Average number of caregivers per class $l$ in childcare type $k$ | # adults | Childcare.gov Supervision: Ratios and Group Sizes (Recommendations), 2025 (33) |

**Table S2:** Summary of contact matrix entries. Formulae used for calculating average number of contacts between index individual of age *i* with contact of age *j* on a given day of care in care type *k* and setting *l* (if relevant, for program care). Parameters are defined in Table S1.

| Childcare type, $k$ | Index | Contact | $C_{i,j}^k$ | $C_{i,j}^{k,l}$ |
| --- | --- | --- | --- | --- |
| None | - | - | 0 | 0 |
| Home care | Young child | Young child | $\left( \frac{p_j^k a_j}{\sum_m p_m^k a_m} \right) \cdot (n^k - 1)$ | - |
| Home care | Young child | Adult | $r_j^k \cdot n_c^k$ | - |
| Home care | Adult | Young child | $p_j^k n^k$ | - |
| Home care | Adult | Adult | 0 | - |
| Program care | Young child | Young child | $\sum_l q_i^{k,l} C_{i,j}^{k,l}$ | $\left( \frac{q_j^{k,l} p_j^k a_j}{\sum_m q_m^{k,l} p_m^k a_m} \right) \cdot (n^{k,l} - 1)$ |
| Childcare center | Young child | Adult | $\sum_l q_i^{k,l} C_{i,j}^{k,l}$ | $r_j^k n_c^{k,l}$ |
| Childcare center | Adult | Young child | $\sum_l C_{i,j}^{k,l}$ | $\left( \frac{q_j^{k,l} p_j^k a_j}{\sum_m q_m^{k,l} p_m^k a_m} \right) \cdot n^{k,l}$ |
| Childcare center | Adult | Adult | $\sum_l C_{i,j}^{k,l}$ | $r_j^k (n_c^{k,l} - 1)$ |

**Table S3:** Characteristics of childcare settings. Attendance statistics, class sizes and caregiver:student ratios from Early Childcare Program Participation study (31) and federal guidelines (33). These values are used to construct contact matrices, 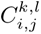.

| Childcare type<br>$k$ | Attendance (days/week)<br>(fraction, $f_i^k$) | Class<br>$l$ | Class size<br>$n^{k,l}$ | Caregiver:Student<br>$\sigma^{k,l}$ |
| --- | --- | --- | --- | --- |
| None | - | - | 0 | 0 |
| Relative home | 3.7 (0.53) | - | 1.8 | 1:1.8 |
| Non-relative home | 3.9 (0.56) | - | 3.9 | 1:3.9 |
| Childcare center | 4.3 (0.62) | Infants, 0-11 months | 6 | 1:3 |
|  |  | Younger toddlers, 1 year | 8 | 1:4 |
|  |  | Older toddlers, 2 years | 8 | 1:4 |
|  |  | Preschool, 3 years | 14 | 1:7 |
|  |  | Pre-Kindergarten, 4 years | 16 | 1:8 |

**Table S4:**
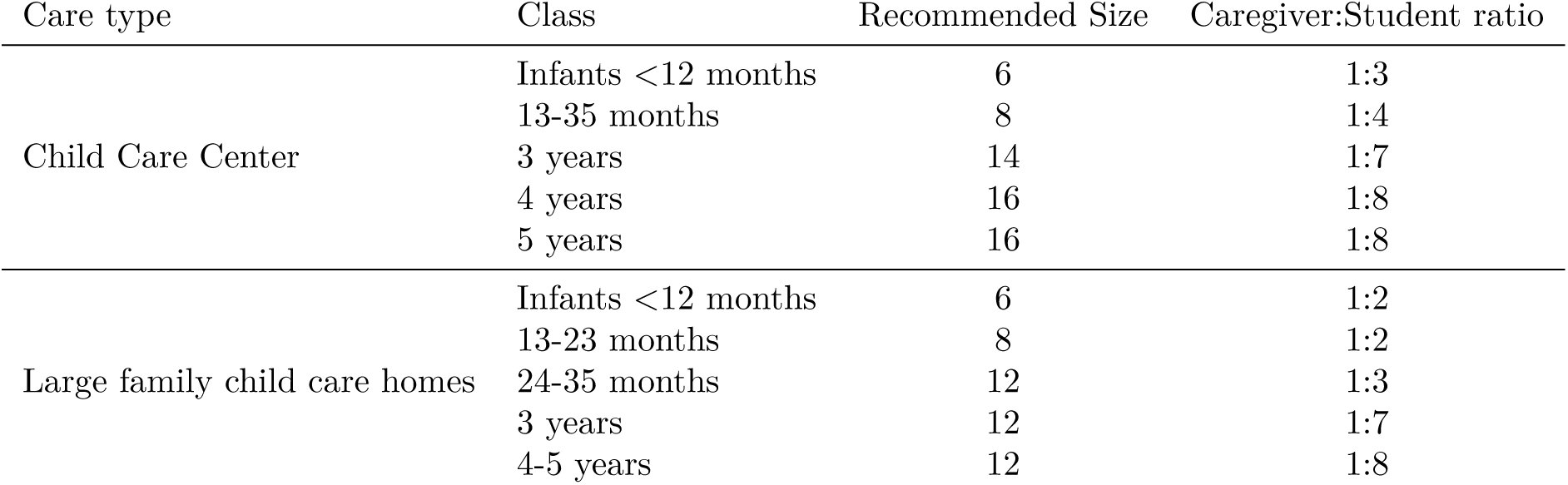
Recommended child size from Office of Child Care. (**33**).

**Table S5:** Questions used from ECPP survey. (**31**). Questions relevant for each care type, and the survey responses allowed within the survey.

| Question | Care type | Responses | ECPP Code |
| --- | --- | --- | --- |
| Is this child now receiving care from a relative other than a parent or guardian on a regular basis, for example, from grandparents, brothers or sisters, or any other relatives? | Relative home | Yes/No | RCNOW |
| About how many days each week does this child receive care from this relative? | Relative home | Integers, 1-7 | RCDAYS |
| How many other children does this relative care for while caring for this child? | Relative home | None, 1-2, 3-5, $\geq 6$ | RCOTCH |
| Is this child now receiving care in your home or another home on a regular basis from someone who is not related to him or her? | Non-relative home | Yes/No | NCNOW |
| About how many days each week does this child receive care from this person? | Non-relative home | Integers, 1-7 | NCDAYS |
| How many other children does this provider care for while caring for this child? | Relative home | None, 1-2, 3-5, $\geq 6$ | NCOTCH |
| Is this child now attending a day care center, preschool, or prekindergarten not in a private home? | Childcare center | Yes/No | CPNNOWX |
| How many days each week does this child go to this program? | Childcare center | Integers, 1-7 | CPDAYS |

